# SARS-CoV-2 IgG antibodies in adolescent students and their teachers in Saxony, Germany (SchoolCoviDD19): persistent low seroprevalence and transmission rates between May and October 2020

**DOI:** 10.1101/2020.07.16.20155143

**Authors:** Jakob P. Armann, Manja Unrath, Carolin Kirsten, Christian Lück, Alexander H. Dalpke, Reinhard Berner

## Abstract

**Introduction:** School closures have been part of the SARS-CoV-2 pandemic control measures in many countries, based on the assumption that children play a similar role in transmitting SARS-CoV-2 as they do in transmitting influenza virus. The contribution of schools in driving the pandemic, however, is still unclear. We therefore performed a SARS-CoV-2 seroprevalence study in students and teachers at two time points in June and October 2020, respectively.

**Methods:** Students grade 8–11 and their teachers in 13 secondary schools in eastern Saxony, Germany, were invited to participate in the SchoolCoviDD19 study. Blood samples were collected in May/June 2020 after the reopening of the schools following the March 2020 lockdown, and again in September/October 2020, 4 weeks after the end of the summer holidays. SARS-CoV-2 IgG were assed using chemiluminescence immunoassay technology and all samples with a positive or equivocal test result were retested with two additional serological tests.

**Results:** 1538 students and 507 teachers were initially enrolled, and 1334 students and 445 teachers completed both study visits. The seroprevalence for SARS-CoV-2 antibodies was 0.6% in May/June and the same in September/October. Even in schools with reported Covid-19 cases before the lockdown of March 13th no clusters could be identified. Of 12 persons with positive serology 5 had a known history of confirmed COVID-19; 23 out of 24 participants with a household history of COVID-91 were seronegative. By using a combination of three different immunoassays we could exclude 16 participants with a positive or equivocal results after initial testing.

**Conclusions:** Schools do not play a crucial role in driving the SARS-CoV-2 pandemic in a low prevalence setting. Transmission in families occurs very infrequently, and the number of unreported cases is low in this age group. These observations do not support school closures as a strategy fighting the pandemic in a low prevalence setting.

## Introduction

Since the identification of the severe acute respiratory syndrome coronavirus 2 (SARS-CoV-2) as the cause of COVID-19 in December 2019 ^1^, the virus spread rapidly around the world, leading to the declaration of a pandemic by the World Health Organization on March 12^th^ 2020. By March 18^th^ 2020, 126 countries—including Germany—had implemented school closures as part of their pandemic control measures, with the number of countries peaking at 194 on April 10^th^ 2020 and more than 90% of the world’s student population being affected at this point^2^.

These actions were mainly based on the assumption that children play a similar role in transmitting SARS-CoV-2 as they do in transmitting influenza during outbreaks, for which evidence exists that school closures reduce the peak of the outbreak^3^. However, there is reason to believe that children play a less significant role in SARS-CoV-2 transmission compared to influenza, making control measures focused on this age group less effective: Most countries—including Germany—report a much lower proportion of cases in children compared to their population size^4–6^. In addition a recent report from Australia could only identify a very limited spread of COVID-19 in primary schools, with no evidence of children infecting teachers ^7^.

However, currently available data is insufficient to rule out that children are as likely as adults to be infected by and to transmit SARS-CoV-2, but simply show little to no symptoms of the disease.

We therefore aimed to quantify the proportion of adolescent schoolchildren and teachers in Saxony, one of the eastern Federal States of Germany, that already have developed antibodies against SARS-CoV-2. Until autumn 2020, in Saxony, the infection rates were comparatively low with 245-laboratory-confirmed SARS-CoV-2 infections per 100,000 inhabitants as of October 13^th^ 2020.

## Methods

### Study Design

After the reopening of the schools in Saxony on May 18^th^, 2020 students grade 8–11 and their teachers in 13 secondary schools in eastern Saxony were invited to participate in the SchoolCoviDD19 study. After teachers, students, and their legal guardians provided informed consent, 5 mL of peripheral venous blood were collected from each individual during visits at each participating school between May 25^th^ and June 30^th^, 2020. In addition, participants were asked to complete a questionnaire on age, household size, previously diagnosed SARS-CoV-2 infections in themselves or their household contacts, comorbidities and regular medication. Students were also asked about regular social contacts outside their household or classroom.

A second visit and repeat blood sampling took place between September 15^th^ and October 13^th^ 2020.

### Approval

The SchoolCoviDD19 study was approved by the Ethics Committee of the Technische Universität (TU) Dresden (BO-EK-156042020) and was registered on July 23^rd^ 2020 and assigned the clinical trial number DRKS00022455.

### Laboratory Analysis

We assessed SARS-CoV-2 IgG antibodies in all samples using a commercially available chemiluminescence immunoassay (CLIA) technology for the quantitative determination of anti-S1 and anti-S2 specific IgG antibodies to SARS-CoV-2 (Diasorin LIAISON® SARS-CoV-2 S1/S2 IgG Assay). Antibody levels > 15.0 AU/ml were considered positive and levels between 12.0 and 15.0 AU/ml were considered equivocal.

All samples with a positive or equivocal LIAISON® test result, as well as all samples from participants with a reported personal or household history of a SARS-CoV-2 infection, were re-tested with two additional serological tests: These were a chemiluminescent microparticle immunoassay (CMIA) intended for the qualitative detection of IgG antibodies to the nucleocapsid protein of SARS-CoV-2 (Abbott Diagnostics® ARCHITECT SARS-CoV-2 IgG) (an index (S/C) of < 1.4 was considered negative whereas one >/= 1.4 was considered positive) and an ELISA detecting IgG against the S1 domain of the SARS-CoV-2 spike protein (Euroimmun® Anti-SARS-CoV-2 ELISA) (a ratio < 0.8 was considered negative, 0.8–1.1 equivocal, > 1.1 positive)

Participants whose positive or equivocal LIAISON® test result could be confirmed by a positive test result in at least one additional serological test were considered having antibodies against SARS-CoV-2.

### Statistical Analysis

Analyses were performed using IBM SPSS 25.0 and Microsoft Excel 2010. Results for continuous variables are presented as medians with interquartile ranges (IQR) and categorical variables as numbers with percentages, unless stated otherwise.

## Results

A total of 1538 students and 507 teachers from 13 different schools participated in the first visit of the study, 1334 students and 445 teachers completed the second visit. Demographic data is shown in Table 1.

**Table 1:**

|  | first study visit (May/June) |  | second study visit (September/October) |  |
| --- | --- | --- | --- | --- |
|  | students | teachers | students | teachers |
| participants | 1538<br>(75.2%) | 507 (24.8%) | 1334 (75.0%) | 445 (25.0%) |
| age (median) | 15 (14–16) | 51 (37–57) | 15 (14–16) | 50 (36–57) |
| female | 802 (52%) | 357 (70%) | 680 (51%) | 313 (70%) |
| household size | 4 (3–5) | 2 (2–4) | 4 (3–5) | 2 (2–4) |
| respiratory symptoms between study visits |  |  | 587 (44%) | 71 (16%) |
| febrile illness between study visits |  |  | 67 (5%) | 4 (0.9%) |

Seroprevalence of SARS-CoV-2 antibodies was 0.6% (12/2045) at the initial visit (May/June) with twelve participants—eleven students and one teacher—having detectable antibodies against SARS-CoV-2 in at least two different assays and thus being considered seropositive. At the follow-up visit (September/October) seroprevalence was 0.7% (12/1779) with still eleven seropositive students and one teacher. Remarkably, one participants who tested positive in the initial sample was no longer positive at the second timepoint and one participant who had equivocal results initially did test positive 3 months later. In 7 out of 13 schools, seropositive participants could be identified, with four seropositive participants in one school as the maximum. The seroprevalence ranged from 0 to 2.2 per individual school.

Of the few participants with a personal history of a SARS-CoV-2 infection, 4/5 were seropositive, with the fifth showing only an equivocal test result in one of the assays. Of all participants with a household history of a SARS-CoV-2 infection, 23/24 were seronegative, with 22/24 showing negative results in all three assays and one showing an equivocal result in only one assay.

During the study period laboratory-confirmed SARS-CoV-2 infections per 100,000 inhabitants in Saxony increased from 139 to 245.

## Discussion

The findings from this unique study in older students and their teachers indicate that the prevalence of IgG antibodies against SARS-CoV-2 was very low after the first wave of the corona pandemic in Germany and during the reopening of the schools in May 2020 and remained low after summer holidays 2020. While this finding is consistent with local surveillance data^8^ that shows a prevalence of PCR-confirmed cases of 0.8%, it clearly indicates that schools did not develop into silent hotspots of SARS-CoV-2 transmission during the first wave of the pandemic and even more importantly after reopening of the schools in May 2020. Even more important is the fact, that there was no increase in seropositivity and infections, respectively, in the four months between May after reopening and October after the summer holidays and the first weeks of back to school in the fall period. It has to be pointed out, however, that the infection rate in Saxony was constantly low during this time period. Nevertheless, the most relevant observation is that infection rates do not increase silently in schools when infection rates in the population are low. Of course, this does not preclude that with increasing infection rates in the population, infection rates in schools may also increase which is an important reminder that the general population has to act prudently in order to keep schools open.

In fact, 5 of the 12 participants with antibodies against SARS-CoV-2 had a personal or household history of COVID-19, yielding a ratio of unidentified to identified cases of 2.4, which is much smaller than that previously assumed by some authors^9^. We could not detect a single cluster of infections in the participating schools, even though at least three schools did have confirmed SARS-CoV-2 cases before the March 13^th^ lockdown in Saxony. This is consistent with findings from the 2003 SARS outbreak ^10,11^, and calls the effectiveness of transmission control measures focused mainly on the student population into question. This is especially relevant since there are clearly described adverse effects of school closures, as loss of education, loss of social contacts and social control, nutritional problems in children who rely on school meals, increases in harm to child welfare in vulnerable populations, as well as economic harm caused by loss to productivity due to parents being forced from work to childcare^12,13^. Additionally, even with school closures in place, social contacts continue as informal child care and non-school gatherings^14^, thereby reducing the potential benefit of school closures further. Our data support this finding this finding since an overwhelming majority of not less than 80% of the participating students in our study reported to have regular social contacts outside their household or classroom.

Close contact with COVID-19 patients—especially in the same household—has been shown to increase viral transmission^15^. However, in our study, only one out of 24 participants with a confirmed SARS-CoV-2 infection in the same household became indeed infected as measured by antibody production. This suggests that either the transmissibility of the virus is lower than previously assumed or that there are certain quarantine and separation measures than can effectively reduce the probability of viral transmission even in close contact situations.

It was reported recently, that SARS-CoV2 spike-reactive CD4+ T cells could be detected in 35% of SARS-CoV2 unexposed healthy blood donors arguing for a certain level of T-cell crossreactivity. Such reactions could arise from exposure to commonly encountered Corona viruses. With children being frequently exposed to common Corona viruses it might be hypothesized that they are less susceptible to SARS-CoV2 infection due to a background of T-cell crossreactivity^16^.

Currently no gold standard serological testing strategy for SARS-CoV-2 exists. Even though immunoassays yield better performance than rapid point-of-care tests^15^ and the targeted SARS-CoV-2 S protein and nucleoprotein show a similarity of less than 30% to endemic betacoronaviruses^17^, false positive results are still a concern, especially in low-prevalence populations and when interpreting results on a personal rather than a population-based level. By using a combination of three different immunoassays and only regarding participants with at least two positive results as seropositive for SARS-CoV-2, we could exclude ten participants with a positive and six with an equivocal initial test by negative confirmatory testing. In our population, a positive predictive value of 42.9% could be observed which was nearby an expected PPV of 45.3% for a prevalence of 0.59% population and the given test characteristics (sensitivity 97.6%, specificity 99.3%). By using this approach, we could reliably identify patients with confirmed seropositivity against SARS-CoV-2 in a low-prevalence population.

## Conclusion

As for now, students and teacher do not seem to play a substantial role in driving the SARS-CoV-2 pandemic in Germany when observing the period after reopening of schools in May as well as after summer holidays until early autumn 2020 before facing the second pandemic wave. Transmission in families appears to occur very infrequently, and the number of unreported cases obviously is low in this age group. For serological testing, a combination of different immunoassays seems to be effective to increase the number of true positive test results.

## Data Availability

The authors confirm that the data supporting the findings of this study are available within the article

## References

1 Zhu N, Zhang D, Wang W, et al. A Novel Coronavirus from Patients with Pneumonia in China, 2019. N Engl J Med 2020; 382: 727–33. https://doi.org/10.1056/NEJMoa2001017.

2 https://plus.google.com/+UNESCO. Education: From disruption to recovery. https://en.unesco.org/covid19/educationresponse (accessed Jul 11, 2020).

3 Cowling BJ, Ali ST, Ng TWY, et al. Impact assessment of non-pharmaceutical interventions against coronavirus disease 2019 and influenza in Hong Kong: an observational study. Lancet Public Health 2020; 5: e279–e288. https://doi.org/10.1016/S2468-2667(20)30090-6.

4 Coronavirus Disease 2019 in Children - United States, February 12-April 2, 2020. MMWR Morb Mortal Wkly Rep 2020; 69: 422–26. https://doi.org/10.15585/mmwr.mm6914e4.

5 COVID-19, Australia: Epidemiology Report 11 (Reporting week to 23:59 AEST 12 April 2020). Commun Dis Intell (2018) 2020; 44. https://doi.org/10.33321/cdi.2020.44.34.

6 Armann JP, Diffloth N, Simon A, et al. Hospital Admission in Children and Adolescents With COVID-19. Dtsch Arztebl Int 2020; 117: 373–74. https://doi.org/10.3238/arztebl.2020.0373.

7 Report: COVID-19 in schools – the experience in NSW | NCIRS. http://ncirs.org.au/covid-19-in-schools (accessed Jul 11, 2020).

8 Zusammenhalt, Sächsisches Staatsministerium für Soziales und Gesellschaftlichen. Infektionsfälle in Sachsen - sachsen.de. https://www.coronavirus.sachsen.de/infektionsfaelle-in-sachsen-4151.html (accessed Jul 12, 2020).

9 Li R, Pei S, Chen B, et al. Substantial undocumented infection facilitates the rapid dissemination of novel coronavirus (SARS-CoV-2). Science 2020; 368: 489–93. https://doi.org/10.1126/science.abb3221.

10 Liao C-M, Chang C-F, Liang H-M. A probabilistic transmission dynamic model to assess indoor airborne infection risks. Risk Anal 2005; 25: 1097–107. https://doi.org/10.1111/j.1539-6924.2005.00663.x.

11 Wong GW, Fok TF. Severe acute respiratory syndrome (SARS) in children. Pediatr Pulmonol Suppl 2004; 26: 69–71. https://doi.org/10.1002/ppul.70056.

12 Jackson C, Mangtani P, Hawker J, Olowokure B, Vynnycky E. The effects of school closures on influenza outbreaks and pandemics: systematic review of simulation studies. PLoS ONE 2014; 9: e97297. https://doi.org/10.1371/journal.pone.0097297.

13 Bin Nafisah S, Alamery AH, Al Nafesa A, Aleid B, Brazanji NA. School closure during novel influenza: A systematic review. J Infect Public Health 2018; 11: 657–61. https://doi.org/10.1016/j.jiph.2018.01.003.

14 Rashid H, Ridda I, King C, et al. Evidence compendium and advice on social distancing and other related measures for response to an influenza pandemic. Paediatr Respir Rev 2015; 16: 119–26. https://doi.org/10.1016/j.prrv.2014.01.003.

15 Pollán M, Pérez-Gómez B, Pastor-Barriuso R, et al. Prevalence of SARS-CoV-2 in Spain (ENE-COVID): a nationwide, population-based seroepidemiological study. The Lancet 2020. https://doi.org/10.1016/S0140-6736(20)31483-5.

16 Braun J, Loyal L, Frentsch M, et al. SARS-CoV-2-reactive T cells in healthy donors and patients with COVID-19. Nature 2020; 587: 270–74. https://doi.org/10.1038/s41586-020-2598-9.

17 Theel ES, Slev P, Wheeler S, Couturier MR, Wong SJ, Kadkhoda K. The Role of Antibody Testing for SARS-CoV-2: Is There One? J Clin Microbiol 2020. https://doi.org/10.1128/JCM.00797-20.

